# Increased serum Neurofilament light chain levels in Parkinson’s Disease patients carrying the p.A53T *SNCA* mutation: Data from the PPMI study

**DOI:** 10.1101/2025.08.28.25334674

**Authors:** Nikolaos Papagiannakis, Christos Koros, Athina-Maria Simitsi, Leonidas Stefanis, The Parkinson’s Progression Markers Initiative

**Author notes:** Corresponding author 1st Department of Neurology, Eginition Hospital, Medical School, National and Kapodistrian, University of Athens, Athens, Greece, 72-74 Vas. Sofias Ave, Athens 11528, Greece. The author list for the Parkinson’s Progression Markers Initiative can be found in the appendix. **Funding:** This research was funded from a grant from the Michael J. Fox Foundation, through the Write Now initiative.

## Abstract

**Introduction:** Serum neurofilament light chain (NfL) levels, a marker of axonal damage, are generally elevated in neurodegenerative conditions, but results in idiopathic Parkinson’s Disease (iPD) have been inconsistent. The p.A53T mutation in the SNCA gene usually leads to a severe, rapidly progressing genetic form of PD.

**Objectives:** Assessing the possible use of serum NfL as a marker of ongoind neurodegeneration in A53T-PD.

**Methods:** Using a propensity score matching based technique with data from the Parkinson’s Progression Markers Initiative, A53T-PD participants were matched, in a 3:1 ratio, to iPD patients and healthy controls by age and disease duration, where applicable.

**Results:** After matching, 18 A53T-PD, 54 iPD, and 54 controls were analyzed. Serum NfL was significantly higher in A53T-PD (9.82 [8.02-18.1] pg/ml) compared to HC (7.54 [5.76-10.1]) or iPD (8.59 [6.86-11.1]) [overall p=0.049].

**Conclusion:** The increase in serum NfL points to a more aggressive neurodegenerative process in A53T-PD compared to iPD. This finding may help design and implement Disease-modifying treatments (DMTs) in this select group of prototypical genetic PD.

## Introduction

Parkinson’s disease (PD) is the most prevalent motor neurodegenerative disease. It is characterized by the loss of dopaminergic neurons in the substantia nigra. Aggregates of misfolded alpha-synuclein in the nervous system, forming Lewy bodies and Lewy neurites, are believed to be crucial in the pathogenesis of PD, although the processes leading to such aggregation are in most cases unknown^1^. In rare instances, pathogenic variants in the SNCA gene, encoding α-synuclein (α-syn), cause familial PD; the first and most studied is the p.A53T missense mutation^2,3^.

Currently, neurofilament light chains (NfL) are recognized as a biomarker of neuroaxonal injury and neurodegeneration in a range of neurodegenerative disorders^4^. NfL is a structural protein found in the axons of neurons, and when axonal damage occurs, NfL is released into cerebrospinal fluid (CSF) and blood, where its levels can be measured to reflect the extent of neuronal damage^5^.

The need for blood-based biomarkers in the diagnosis and monitoring of PD is pressing. Compared to alternative extant methods, testing blood has notable advantages, such as being more accessible for screening and follow-up across various healthcare settings. While blood and CSF biomarkers are still in the early stages of development for PD diagnosis and tracking, they provide valuable insights into the molecular mechanisms potentially driving the disease. Various studies have consistently shown higher NfL levels in the CSF of individuals with idiopathic PD compared to controls ^4^. In contrast, findings in blood, either plasma or serum, have been more inconsistent^6–10^.

The assessment of Nfl levels in genetic PD cohorts is limited, despite these groups offering valuable opportunities to link biomarkers with clinical outcomes and the phenotypic expression of the disease. In one of the few available studies concerning a very small group of patients with the E46K-SNCA mutation, NfL appeared to predict the presence of PD dementia^11^. To our knowledge, no studies have explored serum NfL levels in p. A53T SNCA mutation carriers^3^.

In the dawning age of disease-modifying therapies (DMT), the prompt and precise evaluation of disease severity and progression, through biomarkers such as Nfl, will be crucial for managing patients with PD. This evaluation will aid in the initial assessment, guide the timing for initiating DMT, and facilitate the monitoring of disease progression.

With its vast genetic data and thorough evaluations, the Parkinson’s Disease Progression Marker Initiative (PPMI) database is a great tool for examining clinical and biochemical markers in both healthy controls and a sizable cohort of PD patients, both idiopathic and with genetic mutations^12^. The objective of the current work is to utilize the PPMI data to investigate serum NfL levels in PD participants with the p.A53T point mutation (A53T-PD), as well as in those PD patients who do not have any identified gene variations (idiopathic PD, iPD), and to correlate such levels to disease severity.

## Methods

### Data Acquisition – Sharing

Data used in the preparation of this article were obtained on July 14^th^, 2025 from the Parkinson’s Progression Markers Initiative (PPMI) database (https://www.ppmi-info.org/access-data-specimens/download-data), RRID:SCR_006431. For up-to-date information on the study and data availability, visit http://www.ppmi-info.org. The data used belongs to Tier 1 PPMI data. We have collected except NfL serum levels, basic demographic characteristics (age at visit, age at onset, and disease duration), basic clinical scores (UPDRS parts I to IV) and cognitive tests (Montreal Cognitive Scale, Letter Number Sequencing and Symbol Digit Modalities Test).

### Matching procedure

Each participant – visit pair was considered as one observation for the purposes of matching. For A53T-PD participants, only their earliest visit with NfL data was included, which was the basis for the selection, but there was no such constraint in the other categories.

A propensity score matching based technique was utilized to select the matching HC and iPD participants in a 3:1 ratio with the A53T-PD participants. The propensity score was computed through a logistic regression model with age and age of diagnosis (in the case of iPD only) as independent variables, and the presence of the p.A53T mutation as the dependent variable. Afterwards, a nearest neighbor matching algorithm was used to determine the matching participants. In case a participant was selected more than once with different visits, the visit having the lowest Manhattan distance from the average age and disease duration (for iPD) was kept, while their other visits were removed from consideration. The algorithm was rerun on the new participant–visit set, repeating the aforementioned process until no duplicates remained.

### Statistical Analysis

Statistical analysis was performed using R version 4.5 and sample matching was done with ‘MatchIt’ package. All data were tested for normality using the Shapiro-Wilk test and were determined to be non-normally distributed. We applied the Kruskal-Wallis test, followed by Dunn’s post hoc test for pairwise comparisons, to evaluate differences across the various groups.

Data are presented as median (IQR). Spearman’s rho was used to correlate NfL levels with participants’ age, clinical and cognitive characteristics, with Benjamini-Hochberg correction for multiple comparisons.

The Generalized Extreme Studentized Deviate (ESD) test was used to formally check for outliers in NfL values, after visual inspection of the box plots. A sensitivity analysis was performed after removing the outliers.

The statistical significance limit was set at p = 0.05.

## Results

Serum NfL level data were available for 18 A53T-PD patients for the baseline visit. Consequently, 54 iPD and 54 HC participants were matched to those A53T-PD patients. Their clinical and demographic characteristics are presented in table 1. A53T-PD participants had worse scores on the Montreal Cognitive Assessment and were also quite profoundly hyposmic compared to iPD or controls (table 1). Importantly, all 3 groups were well matched for age at visit, while iPD and A53T-PD had a very similar mean age of onset (Table 1).

**Table 1.** Demographic and clinical characteristics of the study subgroups.

| **Variable** | **Healthy Controls** | **Idiopathic PD** | **A53T-PD** | **p-value** |
| --- | --- | --- | --- | --- |
| Age at Visit (years) | 51.6 (42.5 – 54.3) | 51.02 (46.1 – 54.9) | 50.9 (42 – 54.5) | 0.769 |
| Sex (F/M) | 21/33 | 19/35 | 9/9 | 0.536 |
| Age at Onset (years) | n.a. | 46.0 (39.1 – 49.43) | 46.7 (35.9 – 49.9) | 0.958 |
| Disease Duration (months) | n.a. | 39.2 (28.26 – 64.1) | 48.8 (21.7 – 67.9) | 0.687 |
| MoCA | 29 (27 – 29.8) | 28 (27 – 29) | 26.5 (19.3 – 29) | 0.048 |
| MDS-UPDRS Part III Score | n.a. | 15 (8 – 30.8) | 20 (12 – 28) | 0.234 |
| MDS-UPDRS Part IV Score | n.a. | 0 (0 – 0) | 2 (0 – 3.5) | 0.269 |
| UPSIT | 35 (33 – 36) | 25 (19 – 27) | 12.5 (10 – 18.3) | <0.001 |

Serum NfL levels were increased in A53T-PD participants (9.82 [8.02-18.1] pg/ml) compared to iPD (8.59 [6.86-11.1] pg/ml, p=0.077) or HC (7.54 [5.76-10.1] pg/ml, p=0.008) (overall p=0.049, figure 1). The difference between iPD and HC was not statistically significant (p=0.082).

**Figure 1.**
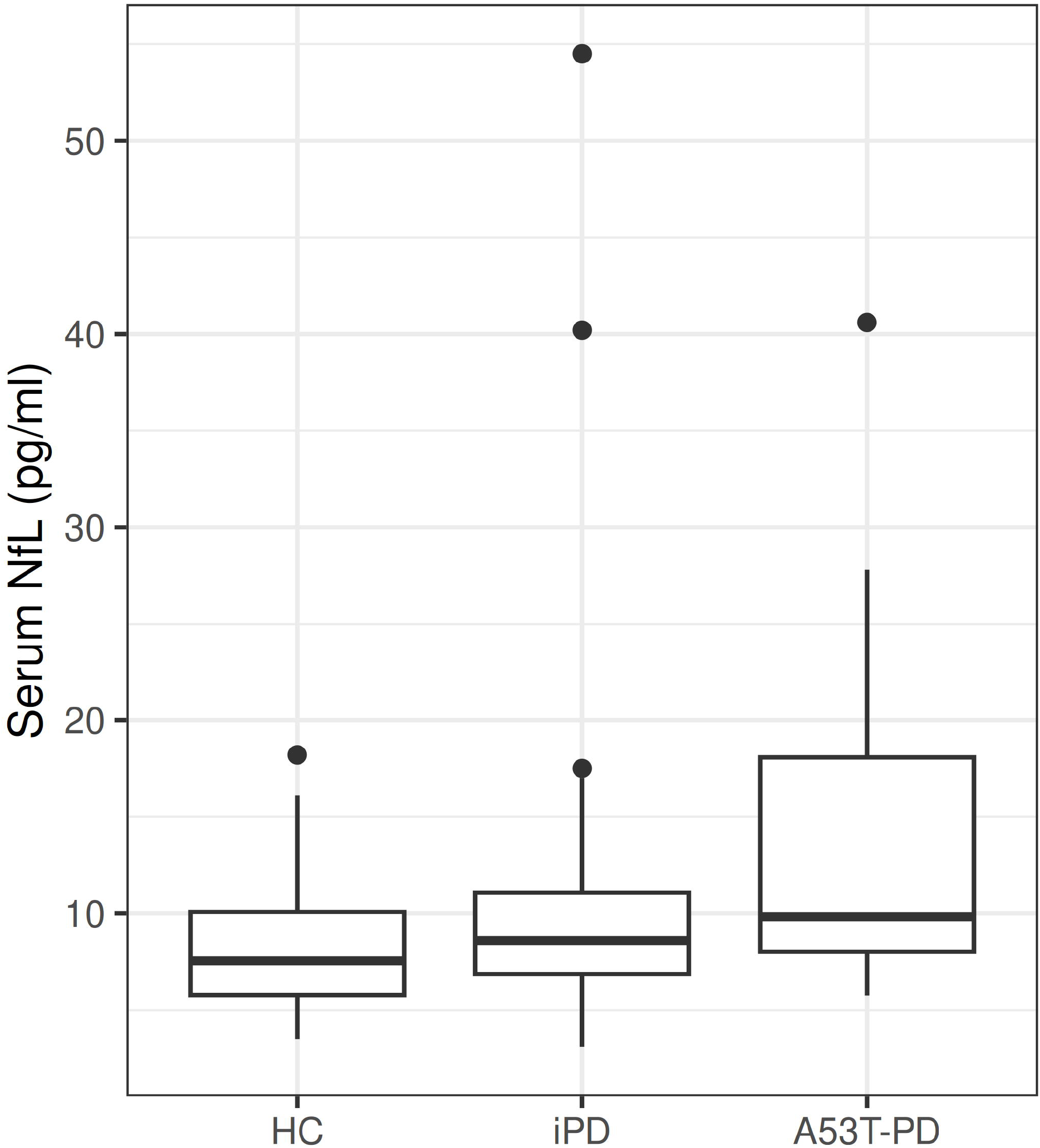

The application of the generalized ESD test revealed that the two most extreme values in the iPD group and the single most extreme value in the A53T-PD group can be identified as outliers (fig. 1). The sensitivity analysis conducted after excluding those three outliers did not reveal any change in the statistical significance, nor did it alter the direction of the results (A53T-PD: 9.68 [8.01-11.1] pg/ml vs iPD: 8.46 [6.85-10.9], p=0.084 or HC: 7.54 [5.76-10.1], p=0.016; p=0.140 between iPD and HC). With the exception of age, no other demographic, clinical or cognitive testing variable was correlated with serum NfL levels in any category. In A53T-PD rho was equal to 0.574 (adjusted p<0.001), while in the iPD and HC groups it was equal to 0.385 and 0.547 respectively (adjusted p=0.060 and p<0.001, supp fig 1).

The data about the longitudinal progression of NfL levels in the A53T-carriers are limited (supp fig 2). They are seem to be mostly stable, with the exception of the participants starting with higher levels, where the increase curve is more steep.

## Discussion

Our results revealed that A53T-PD participants had increased serum NfL levels compared to healthy controls of similar age. Although levels in the A53T-PD group were not significantly higher compared to those of the iPD participants of similar age and disease duration, there was a clear trend. Given the nature of this biomarker, these results point to a more aggressive neurodegenerative process in A53T-PD patients. Overall, patients with A53T-PD exhibit more pronounced clinical and cognitive symptoms^3,13^, which aligns with the observation of increased neurodegeneration, as measured by serum NfL levels. Most studies emphasize the use of serum and CSF NfL levels in PD patients as a predictor of cognitive decline^14,15^. Although it is tempting to attribute the differences in serum NfL levels solely to the increased cognitive burden of A53T-PD participants, we have failed to demonstrate a clear correlation of NfL levels with cognitive scores in the current study.

We were not able to demonstrate a significant correlation of disease duration with Nfl levels in the A53T-PD group (or in the iPD group, for that matter). This would indicate that such levels represent rather a state marker of increased neurodegeneration, regardless of disease stage, at least within the time frame assessed (there were no assessments beyond 4 years’ disease duration). Initial assessments of limited longitudinal data from the PPMI A53T-PD cohort appear to support this, as there is no clear trend of an increase of such levels over time.

We were unable to demonstrate a measurable difference in NfL levels between iPD patients and healthy controls. Although this result is in agreement with some previous studies^6^, the discrepancy with other studies reporting increased levels^7–9^ might lie in the relatively short disease duration of the participants in the present iPD cohort. In most studies, the increased NfL levels were accompanied by cognitive decline or severe motor symptoms^15,16^, neither of which was present here.

NfL levels show a positive correlation with age across all groups, a relationship that has been reported in both healthy individuals and in PD patients^17,18^. This finding emphasizes the importance of the selection of age-matched participants, as age is the single most important confounder.

Serum NfL is strongly correlated with CSF NfL levels, but blood testing is less invasive and more practical for clinical use, supporting the use of serum NfL as a surrogate and convenient early biomarker^16^.

The anticipated influx of new DMTs will increase the urgency of establishing a progression biomarker. In this particular A53T-PD group, serum NfL may serve as a biomarker of ongoing neurodegeneration and as a possible outcome measure in clinical trials of DMTs. Although our preliminary findings do not suggest an increase with increased disease duration, the elevated Nfl levels in this group should respond to successful DMT approaches. In any case, more research on the longitudinal progression of NfL levels in A53T-PD is needed. In addition, these findings may assist in the design and execution of proof of principle clinical trials in this prototypical genetic synucleinopathy.

The main strength of this study is the use of PPMI data. The PPMI database is a very comprehensive longitudinal observational study. Our initial dataset had over 5,000 idiopathic PD participants and approximately 2,000 enrolled healthy controls to select from, enabling us to find participants that had the required demographic characteristics. The main limitation of this study is the small number of A53T-PD cases and the special requirements in their matching with the average iPD patient. It will be interesting to assess whether our results hold in CSF NfL, but there were no available data in the PPMI study for these participants.

In conclusion, serum NfL shows promise as a non-invasive biomarker reflecting neurodegeneration in A53T-PD patients. More extensive longitudinal studies with larger cohorts are essential to validate its utility in tracking disease progression and evaluating therapeutic interventions.

## Data Availability

All data produced in the present study are available upon request to the PPMI core team.

http://www.ppmi-info.org

## Acknowledgments

Statistical analysis codes used to perform the analyses in this article are shared on Zenodo (doi: 10.5281/zenodo.16752152)

PPMI – a public-private partnership – is funded by the Michael J. Fox Foundation for Parkinson’s Research and funding partners, including 4D Pharma, Abbvie, AcureX, Allergan, Amathus Therapeutics, Aligning Science Across Parkinson’s, AskBio, Avid Radiopharmaceuticals, BIAL, BioArctic, Biogen, Biohaven, BioLegend, BlueRock Therapeutics, Bristol-Myers Squibb, Calico Labs, Capsida Biotherapeutics, Celgene, Cerevel Therapeutics, Coave Therapeutics, DaCapo Brainscience, Denali, Edmond J. Safra Foundation, Eli Lilly, Gain Therapeutics, GE HealthCare, Genentech, GSK, Golub Capital, Handl Therapeutics, Insitro, Jazz Pharmaceuticals, Johnson & Johnson Innovative Medicine, Lundbeck, Merck, Meso Scale Discovery, Mission Therapeutics, Neurocrine Biosciences, Neuron23, Neuropore, Pfizer, Piramal, Prevail Therapeutics, Roche, Sanofi, Servier, Sun Pharma Advanced Research Company, Takeda, Teva, UCB, Vanqua Bio, Verily, Voyager Therapeutics, the Weston Family Foundation and Yumanity Therapeutics.

---

**Figure S1.**
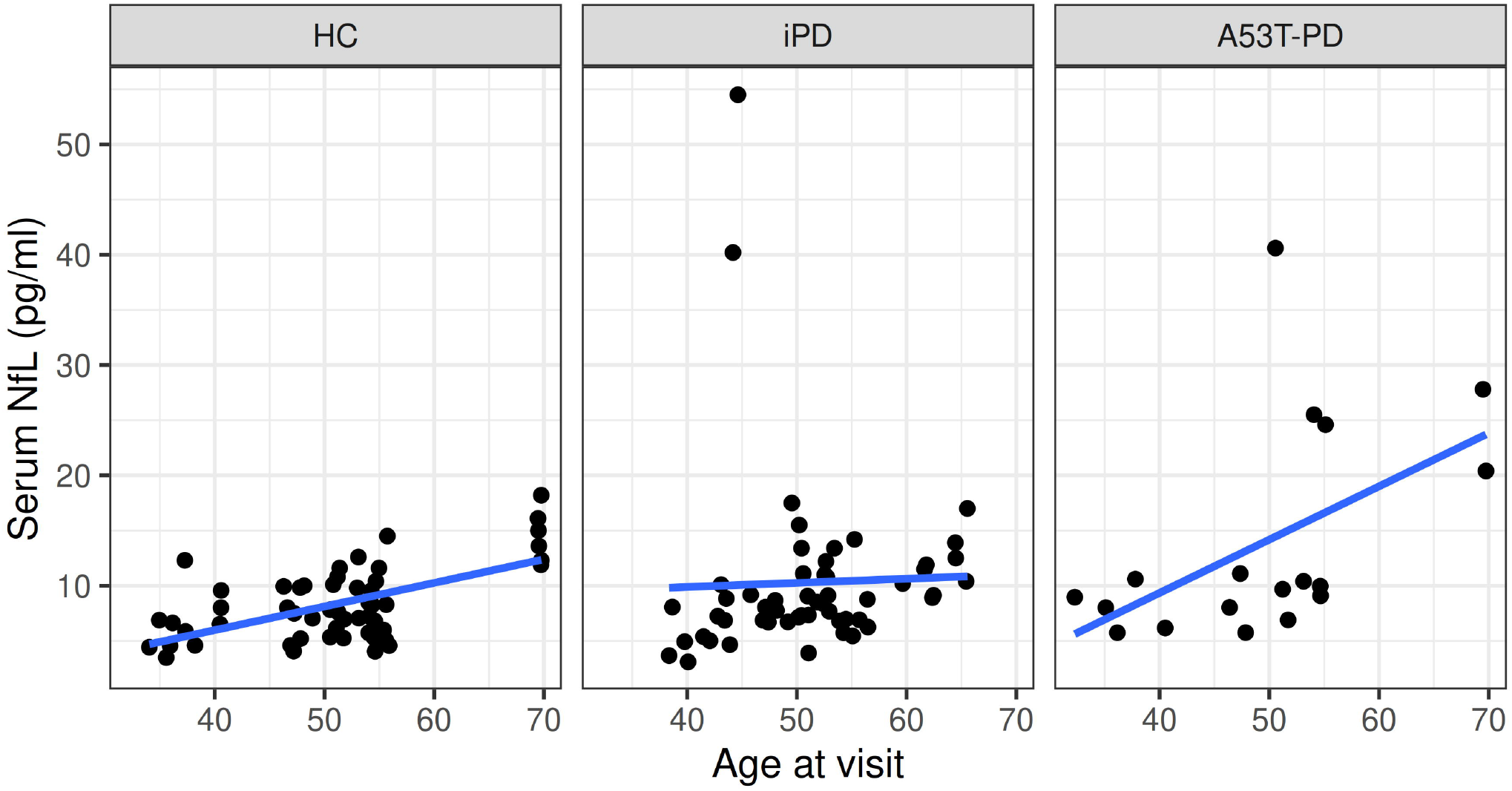

**Figure S2.**
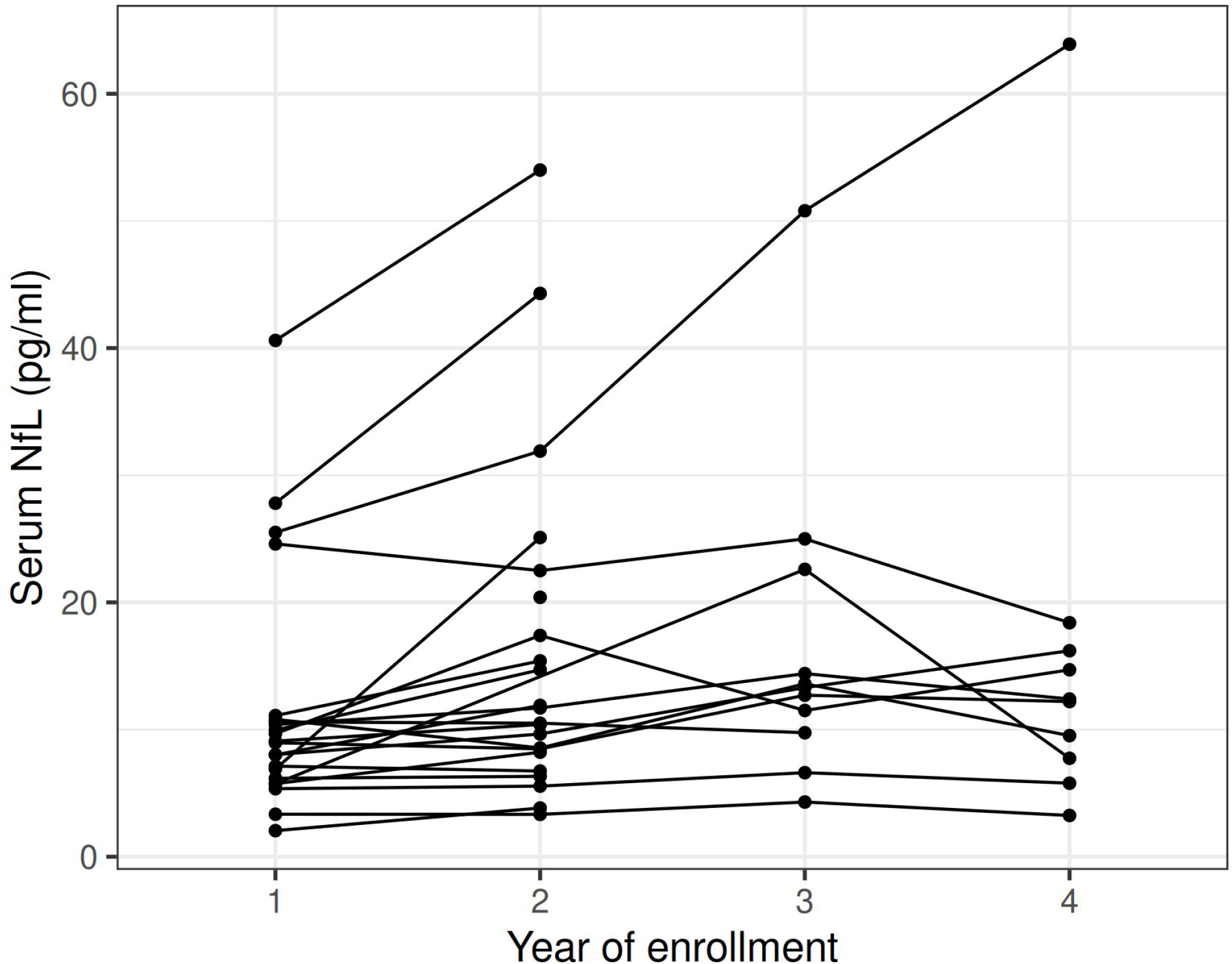

## References

1. Spillantini MG, Schmidt ML, Lee VM, Trojanowski JQ, Jakes R, Goedert M. Alpha-synuclein in Lewy bodies. Nature. 1997;388(6645):839–840. doi:10.1038/42166

2. Papadimitriou D, Antonelou R, Miligkos M, et al. Motor and Nonmotor Features of Carriers of the p.A53T Alpha-Synuclein Mutation: A Longitudinal Study. Mov Disord. 2016;31(8):1226–1230. doi:10.1002/mds.26615

3. Simitsi AM, Sfikas E, Koros C, et al. Motor and nonmotor features of p.A53T alpha-synuclein PD vs idiopathic PD: longitudinal data from the PPMI study. J Neurol. 2025;272(3):203. doi:10.1007/s00415-024-12836-w

4. Bridel C, Van Wieringen WN, Zetterberg H, et al. Diagnostic Value of Cerebrospinal Fluid Neurofilament Light Protein in Neurology: A Systematic Review and Meta-analysis. JAMA Neurol. 2019;76(9):1035. doi:10.1001/jamaneurol.2019.1534

5. Gaetani L, Blennow K, Calabresi P, Di Filippo M, Parnetti L, Zetterberg H. Neurofilament light chain as a biomarker in neurological disorders. J Neurol Neurosurg Psychiatry. 2019;90(8):870–881. doi:10.1136/jnnp-2018-320106

6. Hansson O, Janelidze S, Hall S, et al. Blood-based NfL: A biomarker for differential diagnosis of parkinsonian disorder. Neurology. 2017;88(10):930–937. doi:10.1212/WNL.0000000000003680

7. Ng ASL, Tan YJ, Yong ACW, et al. Utility of plasma Neurofilament light as a diagnostic and prognostic biomarker of the postural instability gait disorder motor subtype in early Parkinson’s disease. Mol Neurodegener. 2020;15(1):33. doi:10.1186/s13024-020-00385-5

8. Aamodt WW, Waligorska T, Shen J, et al. Neurofilament Light Chain as a Biomarker for Cognitive Decline in Parkinson Disease. Mov Disord. 2021;36(12):2945–2950. doi:10.1002/mds.28779

9. Pilotto A, Imarisio A, Conforti F, et al. Plasma NfL, clinical subtypes and motor progression in Parkinson’s disease. Parkinsonism Relat Disord. 2021;87:41–47. doi:10.1016/j.parkreldis.2021.04.016

10. Li Q, Li Z, Han X, et al. A Panel of Plasma Biomarkers for Differential Diagnosis of Parkinsonian Syndromes. Front Neurosci. 2022;16:805953. doi:10.3389/fnins.2022.805953

11. Murueta-Goyena A, Cipriani R, Carmona-Abellán M, et al. Characterization of molecular biomarkers in cerebrospinal fluid and serum of E46K-SNCA mutation carriers. Parkinsonism Relat Disord. 2022;96:29–35. doi:10.1016/j.parkreldis.2022.01.024

12. Marek K, Chowdhury S, Siderowf A, et al. The Parkinson’s progression markers initiative (PPMI) – establishing a PD biomarker cohort. Ann Clin Transl Neurol. 2018;5(12):1460–1477. doi:10.1002/acn3.644

13. Koros C, Stamelou M, Simitsi A, et al. Selective cognitive impairment and hyposmia in p.A53T SNCA PD vs typical PD. Neurology. 2018;90(10):e864–e869. doi:10.1212/wnl.0000000000005063

14. Sheng ZH, Ma LZ, Liu JY, et al. Cerebrospinal fluid neurofilament dynamic profiles predict cognitive progression in individuals with de novo Parkinson’s disease. Front Aging Neurosci. 2022;14:1061096. doi:10.3389/fnagi.2022.1061096

15. Gu L, Zhang P, Gao R, Shu H, Wang P. Predictive value of serum neurofilament light chain for cognitive impairment in Parkinson’s disease. Front Aging Neurosci. 2024;16:1465016. doi:10.3389/fnagi.2024.1465016

16. Liu Y, Dou K, Xue L, Li X, Xie A. Neurofilament light as a biomarker for motor decline in Parkinson’s disease. Front Neurosci. 2022;16:959261. doi:10.3389/fnins.2022.959261

17. Bäckström D, Linder J, Jakobson Mo S, et al. NfL as a biomarker for neurodegeneration and survival in Parkinson disease. Neurology. 2020;95(7). doi:10.1212/WNL.0000000000010084

18. Collins JM, Bindoff AD, Roccati E, Alty JE, Vickers JC, King AE. Does serum neurofilament light help predict accelerated cognitive ageing in unimpaired older adults? Front Neurosci. 2023;17:1237284. doi:10.3389/fnins.2023.1237284

